# Association of Single Nucleotide Polymorphisms with Dyslipidemia in Antiretroviral Exposed HIV Patients in a Ghanaian population

**DOI:** 10.1101/19004812

**Authors:** Christian Obirikorang, Emmanuel Acheampong, Lawrence Quaye, Joseph Yorke, Ernestine Kubi Amos-Abanyie, Priscilla Abena Akyaw, Enoch Odame Anto, Simon Bannison Bani, Evans Adu Asamoah, Emmanuella Nsenbah Batu

## Abstract

Dyslipidemia is a potential complication of long-term usage of antiretroviral therapy (ART) and also known to be associated with genetic factors. The host genetic variants associated with dyslipidemia in HIV patients on ART in Ghana have not been fully explored. The study constituted a total of 289 HIV-infected patients on stable ART for at least a year and 85 aged matched apparently healthy control subjects with no history of HIV and dyslipidemia. Fasting blood was collected into EDTA tube for lipids measurements. Lipid profiles were determined as a measure of dyslipidemia. HIV-infected patients were categorized into two groups; those with dyslipidemia(HIV^-^Dys^+^) (n=90; 31.1%) and without dyslipidemia (n=199; 68.9%) based on the NCEP-ATP III criteria. Four candidate single nucleotide polymorphisms (SNPs) genes (ABCA1-rs2066714, LDLR-rs6511720, APOA5-rs662799 and DSCAML1-rs10892151) were determined. Genotyping was performed on isolated genomic DNA of study participants using PCR followed by a multiplex Ligation Detection Reaction (LDR). The percentage of the population who had the rare homozygote alleles for rs6511720 (T/T), rs2066714 (G/G), and rs10892151 (T/T) and rs662799 (G/G) among HIV^+^Dys^+^ subjects were 5.5%, 14.4%, 6.6% and 10.0%; 2.0% 9.1%, 6.5% and 4.0% among HIV^+^Dys^-^ subjects while 3.5%, 4.7%, 4.7% and 2.4% were observed in HIV^-^Dys^-^ subjects. Statistically significant difference in genotypic prevalence of APOA5 polymorphisms was observed among different groups (p=0.0196). Compared to the AA genotype of the APOA5 polymorphisms, individuals with the rare homozygote genotype [aOR =4.01, 95%CI(1.57-22.39), p=0.004] were significantly more likely to develop dyslipidemia after controlling for age, gender, treatment duration and CD4 counts among the HIV^+^Dys^+^ subjects. There was also a significant associated between GG genotype of ABCA1 and dyslipidemia [aOR =3.29, 95% (1.08 −12.43); p=0.042]. Individuals with the rare homozygote variant (GG) of APOA5 (rs662799) were significantly associated with increased likelihood of developing dyslipidemia [OR =2.24, 95% CI (1.20 −6.83); p=0.0370] holding other variables constant in the HIV^+^Dys^-^ subjects. Our data accentuate the presence of SNPs in four candidate genes and its association with dyslipidemia among HIV patients exposed to ART in the Ghanaian population especially variants in APOA5-rs662799 ABCA1-rs2066714 respectively. These findings provide baseline information that necessitates a pre-symptomatic strategy for monitoring dyslipidemia in ART-treated HIV patients. There is a need for longitudinal studies to validate a comprehensive number of SNPs and its association with dyslipidemia.

## Introduction

Global estimates report 37 million people living with Human Immunodeficiency Virus (HIV), out of which about 26 million reside in Sub-Saharan (SSA) [1]. In Ghana, HIV prevalence among adults aged 15-49 years has declined from about 2.4% in 2013 to 1.6% in 2015 according to the World Bank report [2]. The life expectancy of HIV-infected patients has increased remarkably due to the use of antiretroviral therapy (ART) as a standard of care [3-5]. Unfortunately, long term ART use is associated with a wide spectrum of metabolic disturbances such as lipodystrophy, insulin resistance, and dyslipidemia [6-8]. Dyslipidemia is defined by elevations in total cholesterol, low-density lipoprotein cholesterol (LDL-C), triglycerides and decreased high-density lipoprotein cholesterol (HDL-C).

The prevalence of dyslipidemia is reportedly higher in people living with HIV due to the effect of ART in Ghana [9-11]. The severity of dyslipidemia and the typical pattern of the lipid profile differ between and within the classes of antiretroviral (ARV) agents [12]. Lipid abnormalities have been reported to be frequently associated with HIV-infected individuals receiving protease inhibitors (PIs) and treatment-*naïve* HIV-infected patients, suggesting that HIV infection itself has a metabolic deleterious effect. Such reported side effects are not universal to all individuals on ART and may even vary in individuals with comparable ART, demographic, immunologic and virological characteristics [12-14]. This variability suggests that genetic factors and inherited predispositions may have a significant influence on the incidence of metabolic dysfunction [14, 15]. However, high HDL cholesterol and apolipoprotein A-I (*APOA-I*) have been directly associated with a better immunological outcome [16].

Low-density lipoprotein receptor (LDLR), positioned on chromosome 19p13.2 plays a significant role lipoprotein metabolism by mediating the uptake of cholesterol through the binding and subsequent cellular uptake of apolipoprotein-E and B-constituting lipoproteins. Mutations have been detected in different domains of the LDLR which have distinct effect on LDLR structure and function [17, 18]. ATP-binding cassette A1(ABCA1) plays critical role in reverse cholesterol transport system. Mutation in ABCA1, that encodes this protein, along with genes responsible for their transcription regulation, can lead to abnormality in the metabolism of lipids [19, 20]. Apolipoprotein A5 (APOA5) has been shown to be a key regulator of plasma triglycerides and there are several SNPs associated with the APOA5 gene [21, 22]. Moreover, HIV-infected patients who harbour polymorphisms of the DSCAML1 (Down syndrome cell adhesion molecule like-1) gene exhibit a less favourable lipids profile [23, 24]. Current studies have suggested the relationship between the level of lipids and LDLR, ABCA1 APOA5, and DSCAML1 polymorphisms [15, 25].

Nevertheless, the exact mechanism of dyslipidemia is not fully understood but is most likely multifactorial with the genetic variation being shown to account for about 43-83% of the variability of plasma lipoprotein levels in a normal healthy population [16, 26]. Therefore, from the genetic perspective, HAART-associated hyperlipidemia could be under the influence of various forms of genetic polymorphisms, similar to that in non-HIV adults [27, 28]. Several single nucleotide polymorphisms (SNPs) that could account for a significant portion of the variation of blood lipoprotein concentrations have been identified through recent candidate gene studies and genome-wide association studies (GWAS)[15, 29].

The association between gene polymorphisms that may signal a predisposition to lipid abnormalities and clinical progression of HIV infection has not been thoroughly studied in Africa where the prevalence of HIV is on the increase. SNP prevalence differs by population and at present, the majority of the SNP-associated dyslipidemic studies among HIV patients have come from non-African countries with only a few of these studies emanating from Africa [15, 24, 30]. To the best of our knowledge, no published study has explored the genetic variants and markers associated with dyslipidemia in HIV-infected individuals on HAART in a Ghanaian population. This study, therefore, investigated the distribution of SNPs in four candidate genes that have had significant published lipid associations and their resultant associations with plasma lipid levels in Ghanaian HIV-infected patients on HAART. An understanding of the impact of host genetic factors on the prevalence of dyslipidemia in a cohort of HIV-infected individuals on HAART would promote interventions in the scaling up of treatment regimen.

## Material and Methods

### Study design and subjects

This study comprised HIV-1 infected patients who were on ART regimen for at least one year, with either a protease inhibitor (PI) or non-nucleoside reverse-transcriptase inhibitor (NNRTI) backbone and age-matched HIV seronegative subjects with no history of HIV, dyslipidemia, hypertension, and diabetes The HIV seronegative subjects were referred to as “apparent healthy control” since their nutritional status, diet and quality of life were not assessed in this study. The adjuvant antiretroviral drugs were stavudine, lamivudine, and zidovudine with priorities for inclusion being given to participants who consented to undergo biochemical and genetic testing. Pregnant women, patients being treated with lipid-lowering drugs and those with neurological conditions that prevented them from understanding the concept of the research were excluded from the study.

### Sample size determination

Based on previous report from a study conducted on the burden of dyslipidemia among adults in Ghanaian a population [31] and the lack of knowledge of the frequency of polymorphisms in the population, we assumed an expected proportion of 0.1 for exposure in HIV seronegative subjects, an assumed odds ratio of 2, a confidence interval of 95%, a power of identifying a significant difference between two groups, and 1:3 ratio, a total of 396 subjects were recruited. This comprised of 289 cases HIV seropositive (HIV^+^) subjects and 104 HIV seronegative (HIV^-^) subjects.

### Data collection and biochemical analysis

A structured questionnaire was administered to each patient to obtain demographic information. Details on ARVs, time of diagnosis, duration on ARVs, CD4 counts were obtained from the medical folders of the patients. Fasting blood samples were collected for the analysis of lipid parameters and genomic DNA.

Blood samples were taken after an overnight (12-14 hours) fast into EDTA tubes for biochemical analysis. Fasting lipid panel including total cholesterol (TC), HDL-cholesterol and triglycerides (TG) were measured using Flexor junior (Vital Scientific, Dieren, Netherlands) chemistry autoanalyzer. LDL-C was calculated from the Friedewald’s formula, LDL-C= TC-(HDL-C-TG/2.2). Patient with triglycerides above 4.52 mmol/L was not included in the study since LDL-C was not directly measured and due to the deficit of the Friedewald’s equation, which overestimates LDL-C levels when triglycerides are high. This is to reduce any bias that might affect the relationship found between LDL-C and dyslipidemia. The NCEP-ATP III criteria were used to define dyslipidemia as reduced HDL (<1.03 mmol/L in males; <1.29 mmol/L in females), raised TG ≥1.7 mmol/L, TC >6.2 mmol/L and LDL-C >3.37 mmol/L or specific treatment for such lipid abnormalities [32].

### Anthropometric and hemodynamic measures

Anthropometric measures such as weight and height were performed using an automated weighing scale. Portable height rod stadiometers were used for height measurements to the nearest centimeter. Body mass index (BMI) was defined as weight (kg)/height (m)^2^. Blood pressure was measured using an automated sphygmomanometer (Omron M7 Intelli IT). Three consecutive readings of blood pressure measurements were taken from the patients’ right arm and the mean of two closest values was recorded.

### Single nucleotide polymorphisms selection and Genotyping

The four candidate SNPs (rs2066714, rs6511720, rs662799, and rs10892151) selected for this study have been shown in previous studies to be significantly associated with lipid serum abnormalities following a review of GWAS and PubMed reports of SNPs associated with dyslipidemia among HIV-infected individuals [24, 33-35]. Genomic DNA was isolated from EDTA-collected whole blood samples of study participants using the Qiagen midi kit prep as per manufacturer’s protocol. Genotyping was carried out on isolated genomic DNA of study participants using a multiplex ligation detection reaction (LDR), a sequence-specific genotyping method that has been used efficiently in polymorphism typing and detection of mutations in disease genes [36]. Triplex multiplex reaction assays were setup with products being run on 10% polyacrylamide gel for LDR genotype observation (58 to 90 base pairs). The LDR products differed in sizes of about 8 base pairs for each triplex (Fig. 1). The runs were controlled with Amelogenin XY (AME XY) to determine a successful reaction and gender-confirming for study participants.

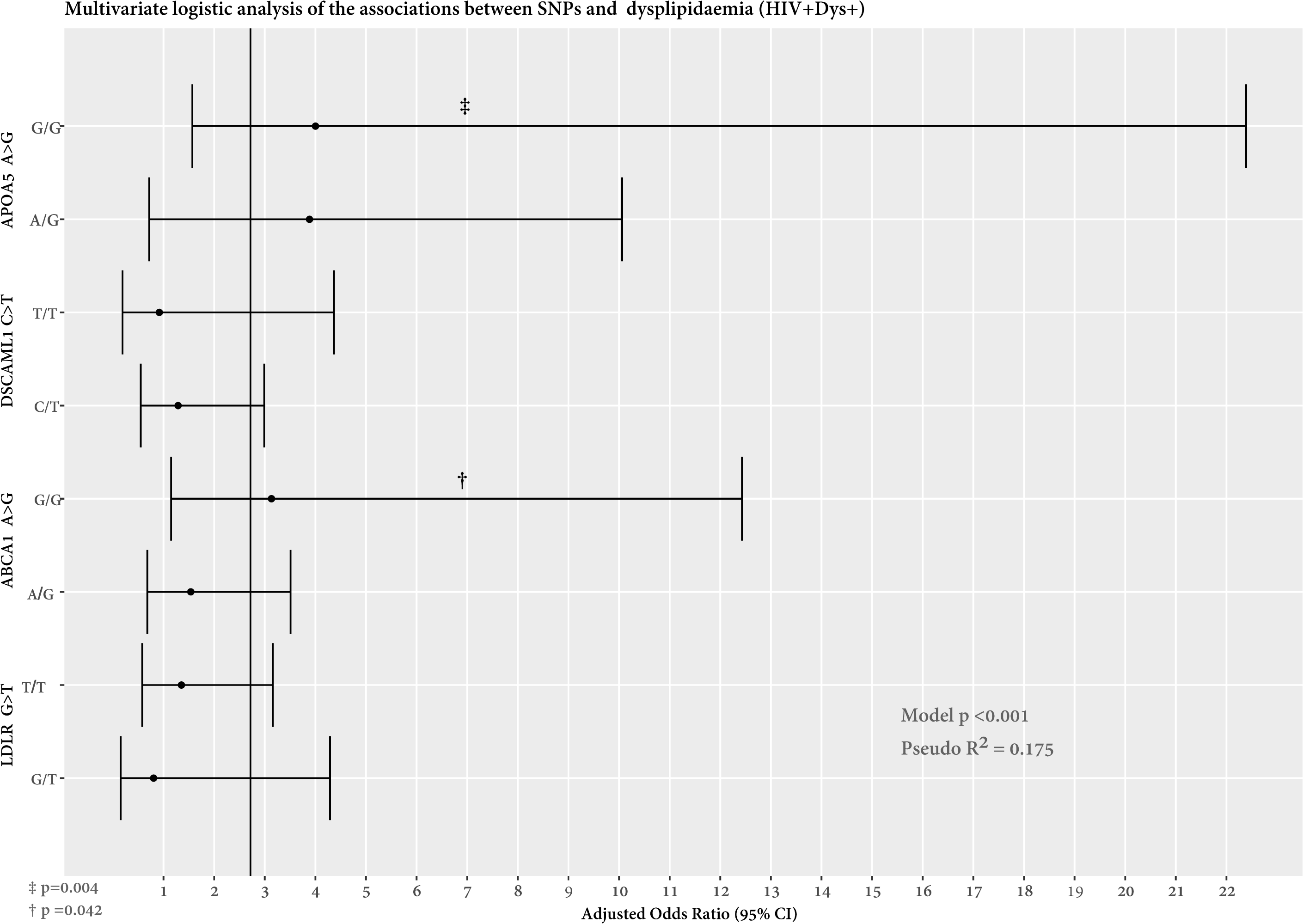

As shown in **Fig. 1**, allele specific LDR products are assigned by color and size following 10% polyacrylamide gel electrophoresis. Each band as shown in the image represents a specific genotype of each allele. At the bottom of the gel are leftover LDR probes from the reaction mix as represented by shared common bands

### Ethical Consideration

The study protocol was approved by the Committee of Human Publication and Research Ethics of the School of Medical Sciences, Kwame Nkrumah University of Science and Technology, Kumasi, Ghana. All participants gave written informed consent and were assured that the information gathered was to be used strictly for research and academic purpose only. In addition, respondents were given the freedom to opt-out at any time they thought they could not continue with the study.

### Data management and statistical Analysis

The NCEP-ATP III criteria were used to defined dyslipidemia among the HIV seropositive and seronegative subjects respectively [32]. The HIV seropositive subjects were categorized into two groups; those with dyslipidemia (HIV^+^Dys^+^) and without dyslipidemia (HIV^+^Dys^-^). Similar categorization was done among the HIV seronegative subjects thus HIV seronegative with dyslipidemia (HIV^-^Dys^+^) and without dyslipidemia (HIV^-^Dys^-^). However, HIV^-^Dys^+^ subjects were excluded from the study since individuals with no history of HIV, dyslipidemia, hypertension, and diabetes were recruited as apparent healthy control. Therefore, nineteen HIV seronegative with dyslipidemia were excluded from the database, allowing for analysis to be performed in 85 apparently healthy control subjects. This was to strengthen the conclusion of the findings reported in this study.

Microsoft Excel software was used to set up a database, and to avoid entry error, the double-entry method was used, and data were analyzed using SPSS version 25 and R program where appropriately. Parametric continuous were analyzed with t-test and expressed as mean ± standard deviation (SD) after checking for normality with Kolmogorov-Smirnov test. ANOVA was used to compare continuous variables from more than two groups with Yates post-test, the chi-square test was to compare differences between more than three groups for categorical variables while Fisher exact was used to assess differences between two groups for categorical variables Allele frequencies were estimated by gene counting. Deviations in the genotype frequencies from the Hardy-Weinberg equilibrium (HWE) were tested using the chi-square (χ^2^) analysis. Initial univariate binary logistic models were performed to determine the association between SNPs and dyslipidemia, followed by adjusted multivariate binary logistic models controlling for age, gender, BMI, CD4 counts and duration of HIV infection. Adjusted multivariate binary logistic models were used also to identify SNPs independently associated with lipid abnormalities holding confounding variables constant. A p-value of less than 0.05 was considered statistically significant.

## Results

**Table 1** shows demographic, hemodynamic indices, lipid parameters, dyslipidemic indices among study subjects based on HIV and dyslipidemia classification. Dyslipidemia was found in 31.1% (90/289) HIV seropositive subjects. No significant difference in age and gender was observed between groups. There were more females than males in all groups (p>0.05. The average duration of treatment among the case subjects was 4 years. There were statistically significant differences in characteristics such as SBP, DBP, weight, height BMI, lipid parameters together with the dyslipidaemic indices (p<05) in all groups. Specifically, HIV^+^ Dys^+/-^ subjects had higher mean levels of SBP, DBP, weight, height, and BMI compared to those of HIV^-^Dys^-^ (p<0.05). Similar trends were observed for the lipid parameters except for HDL-C levels where higher mean levels were recorded for HIV^-^Dys^-^ subjects(1.5±0.3 vs. 1.2±0.5 vs. 0.7±0.3, p<0.0001).

**Table 1:** Comparison of the general characteristics of study participants

| Variables | HIV<sup>+</sup>Dys<sup>+</sup> | HIV<sup>+</sup>Dys<sup>-</sup> | HIV<sup>-</sup>Dys<sup>-</sup> | P-value |
| --- | --- | --- | --- | --- |
|  | n=90 | n=199 | n=85 |  |
| Age (years) | 40.9±10.8 | 39.2±2.0 | 37.8±4.4 | 0.0840 |
| Gender <sup>a</sup> |  |  |  | 0.0920 |
| Female | 73(81.1%) | 157(78.9%) | 59(69.4%) |  |
| Male | 17(18.9%) | 42(21.1%) | 26(30.6%) |  |
| Duration of treatment | 4.3±2.6 | 4.2±2.9 | | 0.1324 |
| CD4 counts | 420.7±28.9 | 410.6±21.2 | | 0.7868 |
| **Blood pressure (mmHg)** |  |  |  |  |
| Systolic | 112.5±13.8 | 114.8±17.6<sup>#</sup> | 110.1±13.1 | 0.0460 |
| Diastolic | 79.9±7.6<sup>¥</sup> | 81.5±8.6<sup>#</sup> | 72.7±12.7 | <0.0001 |
| **Anthropometric indices** |  |  |  |  |
| Weight (kg) | 76.9±13.8<sup>¥</sup> | 76.6±14.5<sup>#</sup> | 65.8±10.2 | <0.0001 |
| Height (m) | 1.58±0.18 | 1.60±0.13<sup>#</sup> | 1.53±0.11 | 0.0003 |
| Body mass index (BMI)<br>(kg/m<sup>2</sup>) | 33.8±2.4<sup>¥</sup> | 31.5±1.1<sup>#</sup> | 28.6±8.7 | 0.0040 |
| **Lipid parameters (mmol/L)** |  |  |  |  |
| Total Cholesterol | 5.2±0.7 | 4.0±0.8 | 4.3±1.3 | <0.0001<sup>‡</sup> |
| Triglycerides | 1.4±0.7 | 1.4±0.7<sup>#</sup> | 1.2±0.5 | 0.0070 |
| HDL-Cholesterol | 0.7±0.3 | 1.2±0.5 | 1.5±0.3 | <0.0001<sup>‡</sup> |
| LDL-Cholesterol | 4.1±0.6<sup>¥</sup> | 2.5±0.8 | 2.5±0.9 | <0.0001 |
| VLDL-Cholesterol | 0.6±0.34<sup>¥</sup> | 0.6±0.35<sup>#</sup> | 0.20±0.08 | <0.0001 |
| Coronary Risk | 10.5±0.31 | 6.2±4.8 | 4.0±0.9 | <0.0001<sup>‡</sup> |
| **Dyslipidaemic indices** |  |  |  |  |
| High triglyceride <sup>a</sup> | 8(8.9%) | 26(13.1%) | 9(10.6%) | 0.0020 |
| High total cholesterol <sup>b</sup> | 10(11.1%) | 4(2.0%) | 0(0.0%) | <0.0001 |
| Low HDL-Cholesterol <sup>a</sup> | 90(100.0%) | 107(53.8%) | 9(10.6%) | <0.0001 |
| High LDL-Cholesterol <sup>b</sup> | 90(100.0%) | 14(7.0%) | 0(0.0%) | <0.0001 |

HIV<sup>+</sup>Dys<sup>+</sup>: HIV seropositive with dyslipidemia, HIV<sup>+</sup>Dys<sup>-</sup>: HIV seropositive without dyslipidemia, HIV<sup>-</sup>Dys<sup>-</sup>: HIV seronegative without dyslipidemia, <sup>a</sup>Chi-square test, <sup>b</sup>Fisher exact test, <sup>#</sup>significant difference between HIV<sup>+</sup>Dys<sup>+</sup> and HIV<sup>+</sup>Dys<sup>-</sup>, <sup>¥</sup> Significant difference between HIV<sup>+</sup>Dys<sup>+</sup> and HIV<sup>-</sup>Dys<sup>-</sup>; <sup>‡</sup> Significant difference between all pairs; Dyslipidemia is defined as the presence of at least one NCEP-ATP III criteria reduced HDL (<1.03 mmol/L in males; <1.29 mmol/L in females), raised TG ≥1.7 mmol/L, TC >6.2 mmol/L and LDL-C >3.37 mmol/L.

**Table 2** shows the frequency distribution of the genotypes and alleles of the four SNPs. The percentage of the population who had the rare homozygote alleles for rs6511720 (T/T), rs2066714 (G/G), and rs10892151 (T/T) and rs662799 (G/G) among HIV^+^Dys^+^ subjects were 5 .5%, 14.4%, 6.6% and 10.0%; 2.0% 9.1%, 6.5% and 4.0% among HIV^+^Dys^-^ subjects while 3.5%, 4.7%, 4.7% and 2.4% were observed in HIV^-^Dys^-^ subjects. Statistically significant differences in allelic frequency of DSCAML1 (p=0.0008)and APOA5 (p=0.0251) among HIV^+^Dys^+^ subjects, ABCA1 (p=0.0010) and DSCAML1 (p=0.0084) among HIV^+^Dys^-^ patients, and none among HIV^-^Dys^-^ subjects. Moreover, chi-square analysis reveals a significant difference in genetic frequency of APOA5 polymorphisms among different groups (p=0.0196).

**Table 2:**
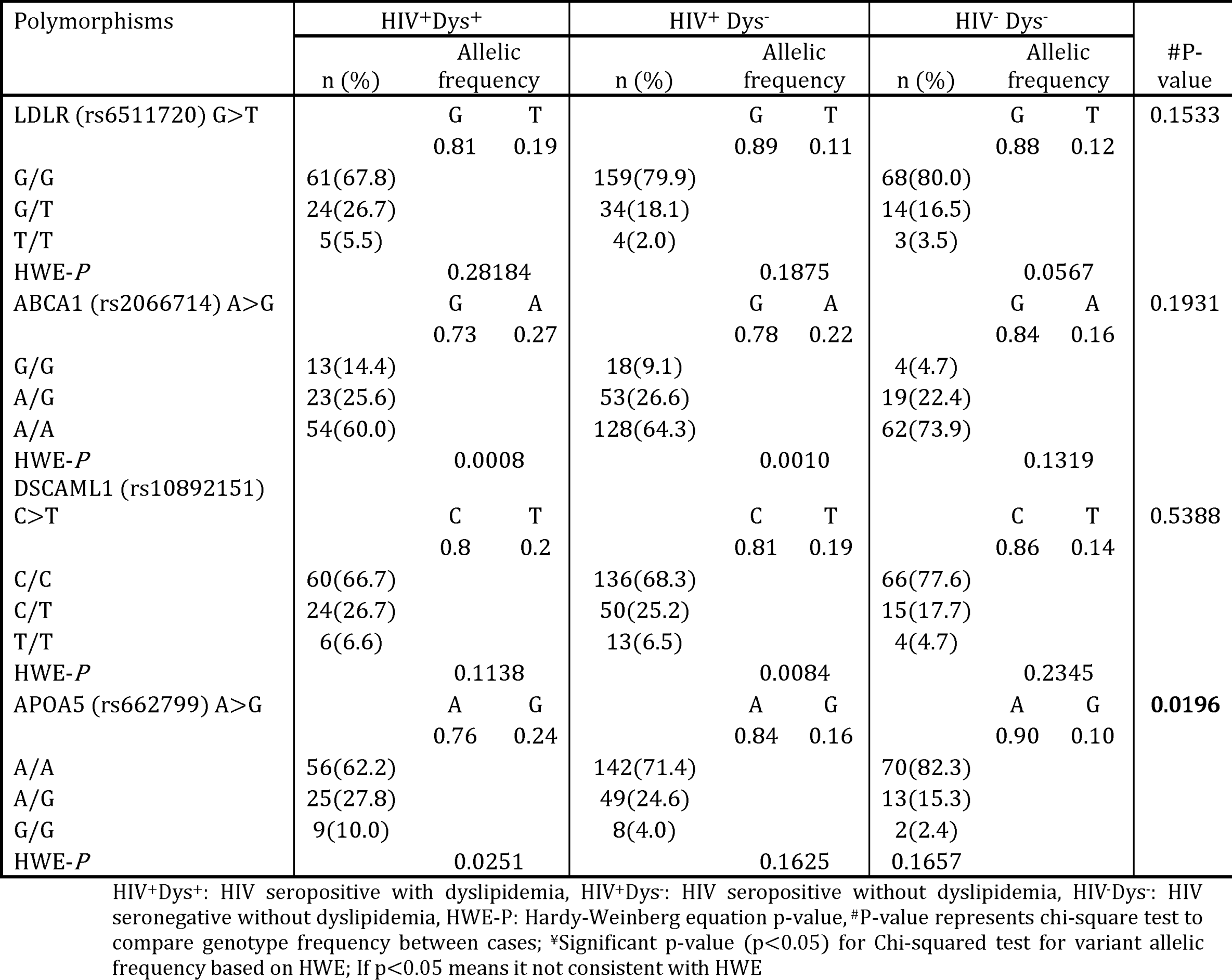
Genotypic and Allelic frequencies of polymorphisms of the Studied Population

### Total Cholesterol (TC)

HIV^+^Dys^+^ subjects with the rare homozygous genotype for *LDLR*, ABAC1 (p<0.0001), LDLR (P=0.0287) DSCAML1 (p=0.0003) and APOA5 (p=0.0151) had a significantly higher level of TC compared to the combined heterozygous and non-carriers genotype. No significant differences were observed in these polymorphisms among the HIV^+^Dys^-^ and HIV^-^Dys^-^ subjects respectively **[Table 3]**.

**Table 3:** Comparison of lipid parameters among study participants based on polymorphisms

| Variables<br>Polymorphisms | HIV<sup>+</sup>Dys<sup>+</sup> |  |  |  | HIV<sup>+</sup>Dys<sup>+</sup> |  |  |  | HIV<sup>-</sup>Dys<sup>-</sup> |  |  |
| --- | --- | --- | --- | --- | --- | --- | --- | --- | --- | --- | --- |
|  | TC<br>(mmol/L) | TG<br>(mmol/L) | HDL-C<br>(mmol/L) | LDL-C<br>(mmol/L) | TC<br>(mmol/L) | TG<br>(mmol/L) | HDL-C<br>(mmol/L) | LDL-C<br>(mmol/L) | TC<br>(mmol/L) | TG<br>(mmol/L) | HDL-C<br>(mmol/L) |
| LDLR (rs6511720)<br>G>T |  |  |  |  |  |  |  |  |  |  |  |
| GG/GT | 5.0±0.6 | 1.4±0.6 | 0.7±0.3 | 4.0±0.5 | 3.9±0.8 | 1.4±0.8 | 1.4±0.5 | 2.4±0.8 | 4.0±0.8 | 1.2±0.5 | 1.5±0.3 |
| T/T | 6.6±1.0 | 2.1±1.8 | 0.6±0.4 | 5.5±0.6 | 4.5±1.4 | 1.6±0.6 | 1.1±0.5 | 2.7±1.2 | 4.3±1.3 | 2.0±0.8 | 1.9±0.5 |
| p-value | <0.0001 | 0.0178 | 0.895 | <0.0001 | 0.1832 | 0.7018 | 0.2125 | 0.4866 | 0.6794 | 0.0017 | 0.0219 |
| ABCA1 (rs2066714)<br>A>G |  |  |  |  |  |  |  |  |  |  |  |
| AA/AG | 5.1±0.7 | 1.3±0.7 | 0.6±0.3 | 4.1±0.1 | 3.8±0.9 | 1.4±0.9 | 1.2±0.6 | 2.4±0.8 | 4.3±1.3 | 1.2±0.3 | 1.5±0.1 |
| G/G | 5.5±0.8 | 1.8±0.7 | 0.8±0.4 | 4.4±0.6 | 4.2±1.1 | 1.4±0.06 | 1.1±0.4 | 2.7±0.9 | 4.5±1.7 | 1.0±0.2 | 1.4±0.2 |
| p-value | 0.0287 | 0.1148 | 0.2213 | 0.0968 | 0.247 | 0.8847 | 0.4001 | 0.3085 | 0.7367 | 0.4425 | 0.7339 |
| DSCAML1<br>(rs10892151)<br>C>T |  |  |  |  |  |  |  |  |  |  |  |
| CC/CT | 4.8±0.5 | 1.2±0.4 | 0.6±0.5 | 4.1±0.6 | 4.0±0.9 | 1.4±0.6 | 1.2±0.5 | 2.3±0.6 | 4.2±1.2 | 1.2±0.5 | 1.4±0.3 |
| T/T | 6.1±1.2 | 2.3±1.5 | 0.9±0.4 | 4.8±0.9 | 4.3±1.0 | 1.9±1.1 | 1.0±0.6 | 2.9±0.6 | 3.8±0.9 | 1.3±0.5 | 1.6±0.3 |
| p-value | 0.0003 | 0.0006 | 0.1036 | 0.0078 | 0.1946 | 0.0133 | 0.3933 | 0.0665 | 0.4101 | 0.7493 | 0.5859 |
| APOA5 (rs662799)<br>A>G |  |  |  |  |  |  |  |  |  |  |  |
| AA/AG | 5.0±0.2 | 1.4±0.7 | 0.6±0.3 | 4.2±0.6 | 4.1±0.7 | 1.5±0.7 | 1.2±0.4 | 2.5±0.8 | 4.3±0.9 | 1.3±0.5 | 1.4±0.3 |
| G/G | 5.6±0.9 | 1.3±0.6 | 0.9±0.2 | 4.5±0.8 | 4.4±1.2 | 1.7±0.9 | 1.0±0.6 | 2.8±0.9 | 4.5±0.7 | 1.5±0.7 | 2.2±0.3 |
| p-value | 0.0151 | 0.7388 | 0.0075 | 0.1017 | 0.1563 | 0.3039 | 0.5685 | 0.2911 | 0.8144 | 0.3248 | 0.0016 |

TC: Total cholesterol, TG: Triglyceride, HDL-C: High density lipoprotein cholesterol, LDL-C: Low density lipoprotein cholesterol, HIV<sup>+</sup>Dys<sup>+</sup>: HIV seropositive with dyslipidemia, HIV<sup>+</sup>Dys<sup>-</sup>: HIV seropositive without dyslipidemia, HIV<sup>-</sup>Dys<sup>-</sup>: HIV seronegative without dyslipidemia, Data is presented as mean±standard deviation, T-test was performed to obtained p-values, p<0.05 significant

### Triglycerides (TG)

Statistically significant differences were observed in TG levels for *DSCAML1* and LDLR polymorphisms between homozygous rare genotype and common genotypes (CC/CT) among HIV^+^Dys^+^ individuals. Similar patterns were observed for DSCAML1 among HIV^+^Dys^-^ patients, and LDLR and APOA5 among HIV^-^Dys^-^ subjects respectively**[Table 3]**.

### HDL Cholesterol (HDL-C)

There were significant differences in HDL-C levels with regards to APOA5 polymorphisms (p=0.0075) in HIV^+^Dys^+^ subjects, and *LDLR* (p=0.0217) and APOA5 A>G polymorphisms (p=0.0016) in HIV^-^Dys^-^ patients respectively. Thus, the combined heterozygous and non-carriers genotype had high levels of HDL-C compared to the homozygous genotypes **[Table 3]**.

### LDL Cholesterol (LDL-C)

There were significant increased LDL-C levels in the rare-allelic subjects for LDLR (p<0.0001) and DSCAML1 (p=0.0078) polymorphisms compared to participants with heterozygous and homogenous genotypes together among the HIV^+^Dys^+^ subjects. No significant differences in the LDL-C levels were noted for any polymorphisms in other groups **[Table 3]**.

Individuals with A/A genotype for ABCA1 (rs2066714)) polymorphisms were significantly more likely to develop dyslipidemia [OR=3.73, 95% CI(1.13-10.89; p=0.0354)] compared with subjects with G/G genotype in HIV individuals with dyslipidemia. The strength of this association was significantly greater in subjects with the rare homozygous [OR=3.67, 95% CI(1.62 – 8.89); p0.0019] and carrier genotypes [OR=5.95, 95% (1.44 – 28.01), p=0.0236] for APOA5 polymorphisms. A similar trend was observed for rare homozygous genotype for APOA5 polymorphisms among HIV^+^Dys^-^ subjects [**Table 4**].

**Table 4:** Univariate logistic analysis of the association of SNPs with the dyslipidemic prevalence

| Variables<br>Polymorphisms | HIV<sup>+</sup>Dys<sup>+</sup> |  | HIV<sup>+</sup>Dys<sup>-</sup> |  |
| --- | --- | --- | --- | --- |
|  | OR (95% CI) | P-value | OR (95% CI) | P-value |
| LDLR (rs6511720) |  |  |  |  |
| G/G | 1 |  | 1 |  |
| G/T | 1.91 (0.89-3.88) | 0.0985 | 1.80(1.01-3.27) | 0.0573 |
| T/T | 1.86(0.46 - 7.20) | 0.4814 | 3.28(0.94-10.91) | 0.1241 |
| ABCA1 (rs2066714) |  |  |  |  |
| G/G | 1 |  | 1 |  |
| A/G | 1.40(0.68 - 2.72) | 0.3745 | 1.02(0.59-1.87) | 0.8825 |
| A/A | 3.73(1.13 - 10.89) | 0.0354 | 1.71(0.79-3.74) | 0.2096 |
| DSCAML1<br>(rs10892151) |  |  |  |  |
| C/C | 1 |  | 1 |  |
| C/T | 1.76(0.83-3.75) | 0.1454 | 1.81(0.98-3.27) | 0.0730 |
| T/T | 1.65(0.41 - 5.36) | 0.5332 | 0.76(0.29-1.96) | 0.6445 |
| APOA5 (rs662799) |  |  |  |  |
| A/A | 1 |  | 1 |  |
| A/G | 3.67(1.62 - 8.89) | 0.0019 | 1.23(0.74-2.38) | 0.3710 |
| G/G | 5.95(1.44 -28.01) | 0.0236 | 3.83(0.34-11.33) | 0.0170 |

HIV<sup>+</sup>Dys<sup>+</sup>: HIV seropositive with dyslipidemia, HIV<sup>+</sup>Dys<sup>-</sup>: HIV seropositive without dyslipidemia, Odds ratio was calculated for each single nucleotide polymorphism using rs6511720-GG, rs2066714-GG, rs10892151-CC, and rs662799-AA genotypes as a referent genotype OR=Odds ratio, CI=Confidence interval. Binary logistic models were performed to obtained SNPs difference for HIV<sup>+</sup>Dys<sup>+</sup> and HIV<sup>+</sup>Dys<sup>-</sup> with references to HIV<sup>+</sup>Dys<sup>-</sup>; p<0.05 significant

**Figures 2** and **3** show the associations between SNPs and dyslipidemia upon multivariate logistic models.

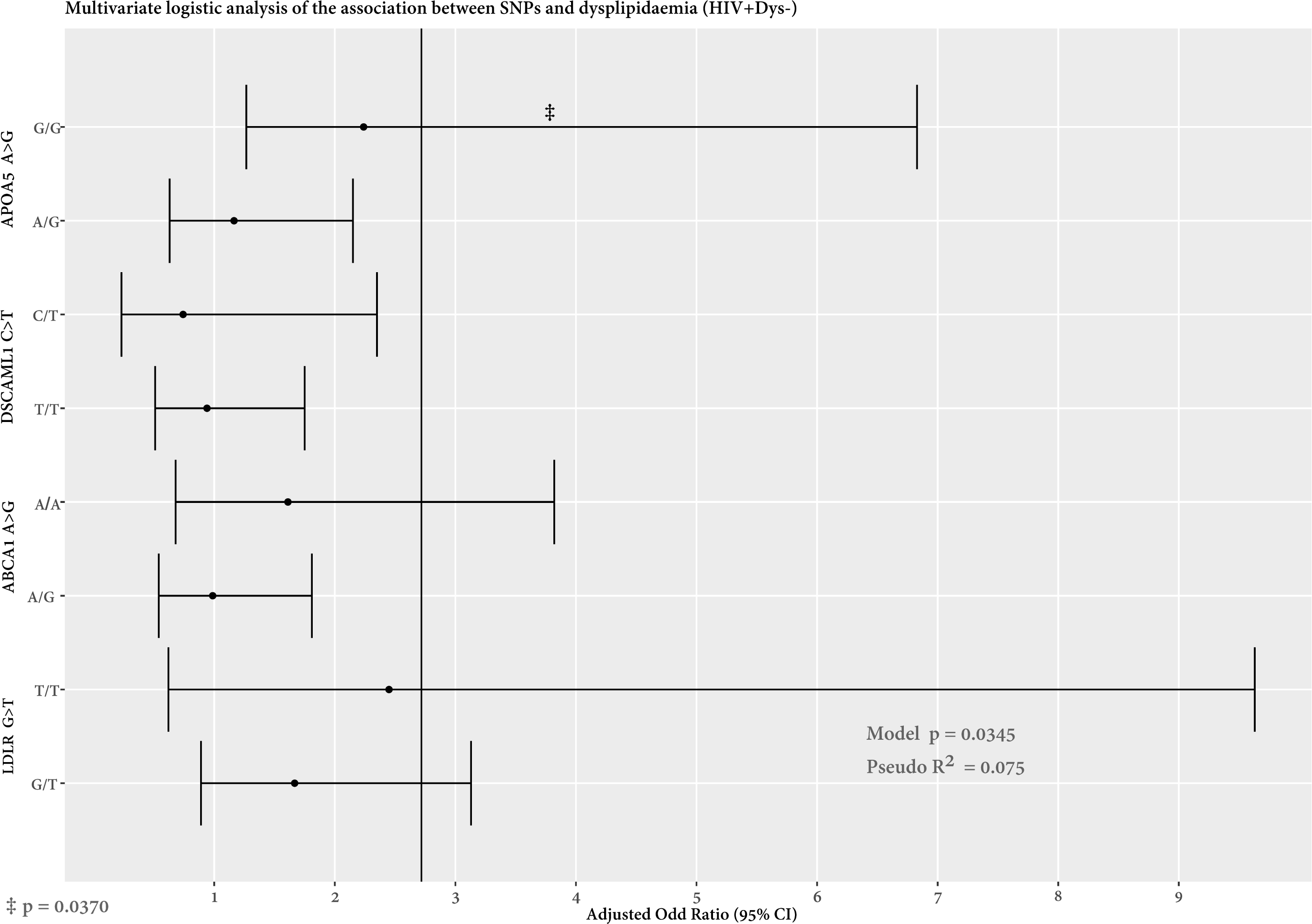

The genetic variants rs6511720-GG, rs2066714-GG, rs10892151-CC, and rs662799-AA genotypes were used as a referent genotype to obtain adds ratio calculations for each single nucleotide polymorphism. OR=Odds ratio, CI=Confidence interval. Binary logistic models were performed to obtained SNPs differences for HIV^+^Dys^+^ and HIV^+^Dys^-^ subjects respectively with reference to HIV^-^Dys^-^ subjects, p<0.05 is considered statistically significant.

Compared to the AA genotype of the APOA5 polymorphisms, individuals with the rare homozygote genotype [aOR =4.01, 95%CI(1.57-22.39), p=0.004] were significantly more likely to develop dyslipidemia after controlling for age, gender, treatment duration and CD4 counts in the HIV^+^Dys^+^ subjects. There was also a significant associated between GG genotype of ABCA1 and dyslipidemia [aOR =3.29, 95% (1.08 −12.43); p=0.042] [Figure 2]. Individuals with the rare homozygote variant (GG) of APOA5 (rs662799) were significantly associated with increased likelihood of developing dyslipidemia [OR =2.24, 95% CI (1.20 - 6.83); p=0.0370] holding other variables constant in the HIV^+^Dys^-^ subjects [**Figure 3**].

**Table 5** shows the association between SNPs with lipid abnormalities after controlling for age, gender, duration of treatment, CD4 counts and BMI respectively. Individuals with the rare homozygous genotype of *APOA5* (G/G) [aOR=5.8(1.8-44.9), p=0.0009], *ABCA1* (G/G) [aOR=10.6(1.3-88.6), p=0.028] and *LDLR* (rs6511720) G>T [aOR=21.2(7.6 −49.4), p<0.0001) were more likely to have high levels of TC levels. Moreover, subjects with T/T genotype of *APOA5* (G/G) polymorphism were associated with increased levels of LDL-C [aOR=2.2(1.4-6.0), p=0.014] among the HIV^+^Dys^+^ subjects. Furthermore, the homozygous genotype of LDLR (T/T) [aOR=11.0(1.9-63.6); p = 0.007)) were significantly associated with high TC levels.

**S1. Table 1** shows the demographic, clinical and metabolic characteristics of participants stratified on the type of ART (NNRTI vs. PI). There were 3.5% (10/289) on PI-based treatment whiles 279 (96.4%) were on NNRTI-based treatment. No statistically significant association was observed between NNRTI AND PI-based treatment in relation to age (p=0.2569), gender (p=0.6929), duration of treatment (p=0.2092), systolic (p=0.7638) and diastolic pressure (p=0.0865), weight (p=0.2122), height (p=0.7221) and BMI (0.6825) respectively. Seventy-five (15.6%) patients had hypertriglyceridemia, 24.9% had hypercholesterolemia and 71.9% had low HDL-C in relation to metabolic parameters. PI-based subjects had significantly higher levels of total cholesterol (6.09±0.56 vs. 4.34±0.96, p=0.0001) and LDL cholesterol (4.60±0.55 vs. 2.95±1.16, p=0.0001) and triglycerides (2.61±0.65 vs. 1.43±0.69, p=0.0001) compared to NNRTI-based subjects.

**Table 5:** Association of SNPs with individual lipid abnormalities

| Variables | High TC<br>(mmol/L) | High TG<br>(mmol/L) | Low HDL-C<br>(mmol/L) | High-LDL-C<br>(mmol/L) |
| --- | --- | --- | --- | --- |
|  | aOR (95% CI) | aOR (95% CI) | aOR (95% CI) | aOR (95% CI) |
| HIV<sup>+</sup>Dys<sup>+</sup> |  |  |  |  |
| LDLR (rs6511720) T/T | 21.2 (7.6 -49.4)<br>*** | 0.9 (0.1 -1.0) | 0.9 (0.2 - 3.9) | 2.7 (0.7 - 10.6) |
| ABCA1 (rs2066714) G/G | 10.6 (1.3 - 88.6) * | 1.8 (0.6 - 5.3) | 1.3 (0.5 - 3.2) | 1.7 (0.8 - 3.9) |
| DSCAML1 (rs10892151)<br>T/T | 2.3 (0.3 - 17.2) | 1.7 (0.5 - 6.2) | 0.8 (0.3 - 4.8) | 0.9 (0.3 - 2.8) |
| APOA5 (rs662799) G/G | 5.8 (1.8 - 44.9) ** | 0.9 (0.2 -4.6) | 1.4 (0.4 - 4.8) | 2.2 (1.4 - 6.0) * |
| HIV<sup>+</sup>Dys<sup>-</sup> |  |  |  |  |
| LDLR (rs6511720) T/T | 11.0 (1.9 - 63.6)<br>** | n/a | 1.1 (0.2 - 4.5) | 1.6 (0.3 - 7.5) |
| ABCA1 (rs2066714) G/G | 4.2 (0.8 - 23.1) | 1.9 (0.4 - 9.4) | 2.8 (0.9 - 9.2) | 2.9 (0.8 - 10.3) |
| DSCAML1 (rs10892151)<br>T/T | 1.8 (0.2 - 14.0) | 0.9 (0.06 - 12.2) | 0.6 (0.1 - 2.6) | 0.7 (0.2 - 3.4) |
| APOA5 (rs662799) G/G | 4.3 (0.8 - 24.5) | 2.8 (0.4 - 20.6) | 1.0(0.3 -3.7) | 3.6 (0.7 - 19.1) ** |

HIV<sup>+</sup>Dys<sup>+</sup>: HIV seropositive with dyslipidemia, HIV<sup>+</sup>Dys<sup>-</sup>: HIV seropositive without dyslipidemia aOR=Adjusted odds ratio, TC =Total cholesterol, TG= Triglycerides, HDL-C=High Density Lipoprotein Cholesterol, LDL-C=Low Density Lipoprotein Cholesterol, , Odds ratio was calculated for rare recessive polymorphism of rs6511720, rs2066714, rs10892151 and rs662799 genotypes using a combination of carriers and homozygotes as a referent genotype, OR=Odds ratio, CI=Confidence interval. Binary logistic models were performed to obtained SNPs difference for HIV<sup>+</sup>Dys<sup>+</sup> and HIV<sup>+</sup>Dys<sup>-</sup> with references to HIV<sup>+</sup>Dys<sup>-</sup>; \*p<0.05, \*\*p<0.001, \*\*\*p<0.005, p<0.05=Statistically Significant

## Discussion

Long term usage of ART has been implicated in several metabolic effects including dyslipidemia. However, this side effect varies among individuals on ART with comparable clinical and demographic characteristics. As such, inherited predispositions and genetic factors have been implicated in the cause of this metabolic alteration. The present study, therefore, assessed the prevalence of SNPs and its association with dyslipidemia in HIV patients on ART in Ghana. Four candidate SNPs (rs2066714, rs6511720, rs662799, and rs10892151) reviewed from PubMed were analyzed for associations with dyslipidemia among HIV-infected individuals.

Several studies have shown an association between dyslipidemia and HIV patients on ART [37, 38]. In the present study, the prevalence rate of dyslipidemia among HIV patients on ART was 31.1% with low HDL-C and high TC levels being the commonest lipid abnormalities. The observed prevalence of dyslipidemia is comparable to the range of reports from previous studies by Obirikorang et al.[9] and Ngala et al. [10] in Ghana and Kodogo et al. [10] in Zimbabwe. However, it is lower compared to 78.9% prevalence rate reported by Limas et al. [39] in a cross-sectional study among Brazilian HIV individuals on ART. Furthermore, our findings of low HDL-C and high total cholesterol and LDL-C among HIV patients on ART are consistent with other studies [40, 41]. Results from the present study revealed that subjects who received PI-based treatment had significantly increased levels of total cholesterol, LDL cholesterol and triglycerides compared to non-PI-based subjects. This is consistent with several case reports [42, 43] and cross-sectional studies [44, 45] which have reported that PI exposures are associated with hypercholesterolemia and hypertriglyceridemia.

Our study demonstrated that the presence of the homozygous recessive/mutant genes for *LDLR, ABCA1, DSCAML1* and *APOA5* genotypes among our study participants were very low [Table 2]. Similar low frequencies in APOA5 and LDLR homozygotes mutant genes were reported by Lazzaretti et al. [15] in a cross-sectional study among HIV-infected patients on ART in the Brazilians population. Genome-wide associations studies have identified LDL-R SNP rs6511720 (G>T), which is located in intron-1 of the gene, to be associated with lower plasma levels of LDL-C and a lower risk of CHD [35]. Data from the GLGC consortium suggested that *LDLR* rs6511720 minor allele is prevalent in about 10% of the population and have established that the allele is protective, being associated with lower levels of LDL-C [46].

Notwithstanding that, the frequency of recessive genes was low in this study population, the prevalence of the homozygous genotypes for the minor alleles of LDLR, DSCAML1, ABACA1, and APOA5 SNPs was higher in the HIV^+^Dys^+/-^ subjects compared to the HIV^-^Dys^-^ subjects. All but APOA5A SNPs did not record any significant genotypic prevalence. These findings are consistent with previous reports by Wang et al., [19] and Aragones et al. [24] who did not find any significant differences in their studies with respect to prevalence of the DSCAML1 and ABCA1 genotypes between cases and control subjects. Our findings demonstrated that the allelic frequencies were in equilibrium with the Hardy-Weinberg equation among the HIV sero-negative subjects without dyslipidemia, nonetheless, genotypic frequencies of ABCA1 and APOA5 polymorphisms were not consistent with Hardy-Weinberg equation among the HIV^+^Dys^+^ subjects, as well as ABCA1 and DSCAML1 polymorphisms among the HIV^+^Dys^-^ subjects. The low frequencies of the homozygotes of the rare alleles of theses SNPS contributed to the observed deviations.

A longitudinal study conducted by Rotgar et al. [47] validated the contribution of 42 SNPs to dyslipidemia among HIV-infected population treated with ART. The authors reported that the degree of the contribution of SNPs and ART to dyslipidemia are similar and therefore genetic information should be considered in addition to the dyslipidemic effect of ART agents[47]. Results from the present study revealed that individuals with rs6511720 T/T genotype recorded increased levels of TC, TG, and LDL-C compared to non-carriers and heterozygous combined rs6511720 GG/GT [Table 3]. Further multivariate logistic model analysis showed a strong association of LDLR polymorphisms with high total cholesterol holding all other confounding variables constants. A study by Lazzaretti et al. [15] reported that *LDLR* intron 19G>T (rs6511720) did not contribute to the plasma lipid levels in their dataset, and hence may reflect a limited effect of this SNPs in HIV-infected patients. The inconsistency could be due to different geographical settings and thereby calls for more research in these SNPs among African descendants.

In this study, ABCA1 (rs2066714) G/G genotype was significantly associated a higher probability of developing dyslipidemia. Furthermore, subjects with the homozygotes of the rare alleles of ABCA1 have increased levels of TC compared to the common allele (AA/AG) genotypes. As a further matter, subjects carrying the minor allelic variant of APOA5 were 2 times more likely to have low levels of HDL cholesterol among the HIV population in the present study though no statistical significance was observed. These observations are in congruent with previous reports demonstrated in literature demon ABCA1 is associated with familial HDL deficiency. We should mention here that low HDL-C was one of the commonest lipid abnormalities observed in this study. Moreover, ABCA1 mediates the efflux of cellular cholesterol and phospholipids unto ApoA-I and thereby plays a central role in regulating cellular cholesterol homeostasis, and forming HDL[48, 49].

*DSCAML1* (rs10892151) SNP in this study had increased levels of TC, TG, and LDL-c in T/T genotype individuals compared with the other genotypes. Further analysis indicated that individuals with the homozygous minor recessive gene (carriers) (T/T) were more likely to have increased levels of TC and LDL-C. These observed findings are in parallel with previous reports by Aragones et al. [24] who reported a strong association between the expression of the rs10892151 T allelic variant and dyslipidemia, mostly hypertriglyceridemia and depressed HDL-cholesterol levels. In contrast, Pollin et al., [23] observed that rs10892151 T carriers had lower fasting and postprandial serum triglycerides values than non-carriers, and they found a linkage disequilibrium with an APOC3 null mutation, which was likely the result of a founder effect in their high-fat feeding intervention study. Evidence provided in the literature shows that carriers of this null mutation have low circulating apolipoprotein (Apo) C-III levels and reduced fasting and post-prandial triglyceride concentrations [23], which is likely due to the well-established function of Apo C-III as an inhibitor of lipoprotein lipase[50].

This current study showed that carriers of the minor allelic variant of the *APOA5* (rs662799) A>G SNP gene, were 4 times more likely to developed dyslipidemia. Moreover, individuals with homozygotes (G/G) have significantly increased levels of TC and LDL cholesterol compared to wild type and heterozygote combined (AA/AG). Previous studies have found rs662799 to be associated with elevated plasma triglyceride levels as well as HDL-C and total cholesterol [21]. Two polymorphisms in the *APOA5* gene, −1131T>C and S19W (56C>G), have already been shown to be associated with elevated triglyceride levels in different populations[51, 52]. This study, however, considered the *APOA5* (rs662799) A>G variant and found that HIV individuals with at least one G allele had higher TC and LDL cholesterol levels.

Lazzaretti et al. [15] reported that *APOA5* −1131T>C (rs662799) was associated with plasma triglycerides (TG) and low-density lipoprotein cholesterol levels (LDL-C) as well as high-density-lipoprotein cholesterol levels. To the best of our knowledge, this study is the first to investigate the *APOA5* (rs662799) A>G variant in HIV patients in a Ghanaian population. Another study by Echeverria et al. [53] reported that polymorphisms in genes associated with the development of atherogenic dyslipidemia, especially variants in the *APOA5* gene, can influence the circulating CD4 T-cell levels in chronically HIV-infected patients. Although a previous study has reported the effect of APOA5 on CD4 levels, individuals with the minor allelic variant of *APOA5* (rs662799) A>G have a significant likelihood of developing dyslipidemia in HIV subjects on ART after controlling for CD4 counts as well as age, gender, duration of treatment and BMI.

This study has strength in it possibly being the first study to assess the prevalence of SNPs and its association with dyslipidemia in the Ghanaian population as a proof of concept. Although previous reports have shown an uneven continuous rate of the incidence of HIV in the Ghanaian population with more than 60% of people living with HIV are females [54], the proportion of female in the HIV study population was much higher than males which could affect the generalizations of our findings to the HIV population in Ghana. In addition, our study is limit by the fact that the number of investigated polymorphisms is not comprehensive, notwithstanding, this is a baseline study for future exploratory analysis of SNPs and its association with dyslipidemia among HIV patients on ART in Ghana.

## Conclusion

This study has highlighted the evidence that SNPs in four candidate genes are present in HIV patients exposed to ART in the Ghanaian population. Dyslipidemia remains prevalent among HIV patients. SNPs were found to associate with dyslipidemia especially variants in APOA5-rs662799 ABCA1-rs2066714 respectively. This finding provides baseline information that necessitates a pre-symptomatic strategy for monitoring dyslipidemia in ART-treated HIV patients Findings from this study should be validated in a longitudinal case-control study considering the disease and its therapeutic implications. The candidate SNP if validated will help in serving as potential biomarkers to detect individuals at risk for dyslipidemia.

## Data Availability

The data used to support the findings of this study are part of an ongoing project and are available upon reasonable request

## Acknowledgment

The authors thank Prof. David T. Burke and Jodi Wilkowski of the Department of Human Genetics, University of Michigan Medical School, Michigan, USA for immense help. We also wish to express our profound gratitude to all HIV patients who actively participated in this study.

## Data Availability

The data used to support the findings of this study are part of an ongoing project and are available upon request.

